# Genetic subtypes predict multiple sclerosis severity and response to treatment

**DOI:** 10.1101/2025.04.15.25325853

**Authors:** Karim L Kreft, Nienke J. Mekkes, Emeka Uzochukwu, Sam Loveless, Ray Wynford-Thomas, Katharine E. Harding, Mark Wardle, Peter Holmans, J William L Brown, Michael Lawton, Emma C Tallantyre, Inge R. Holtman, Neil P Robertson

## Abstract

**Background:** Predicting response to treatment and risk of long-term disability in multiple sclerosis (MS) is challenging. In other disease areas, combining genetic risk variants enabled detection of relevant clinical endophenotypes associated with important outcomes, but this has never been applied to MS.

**Methods:** We applied unsupervised hierarchical clustering to genomic risk scores of two cohorts (the prospective cohort study of 1455 Welsh MS patients was used as the discovery cohort and replication was performed in a multi-centre post-mortem Netherlands Brain Bank cohort of 272 MS patients) to predict relevant disease outcomes using survival analysis for time to disability milestones (expanded disability status scale, EDSS), and ANOVA to compare linear clinical outcomes.

**Results:** Three genomic clusters were identified, in each cluster patients had similar genetic profiles. Baseline demographic characteristics were similar between clusters. Welsh patients in cluster 1 attained key disability milestones later, reaching EDSS6, 6 years later (p=0.003) and EDSS8, 13 years later (p=0.02) than those in clusters 2 and 3. Time to EDSS6 was also significantly longer for patients in cluster 1 versus cluster 2 in the NBB-MS cohort (6 years, p=0.04). Genomic clustering is an independent predictor for disease progression compared with well-validated risk factors (Hazard ratio for time to EDSS6 1.3-2.0, all p<0.05). Welsh patients in cluster 2 and 3 also had a significantly greater annual increase in T2 lesion load on serial MR imaging (p=0.04). In cluster 2, patients who had received MS disease modifying treatments (DMT) had a longer time to EDSS6 (p=0.003) compared to those that had received no DMTs, whereas no differences were observed in either cluster 1 or cluster 3. In the NBB-MS cohort, we also observed differences in symptomatology, including earlier development of swallowing problems (p=0.02) or muscle spasticity (p= 0.0008) in cluster 2 patients.

**Conclusion:** This study demonstrates that unsupervised genetic clustering has utility to detect clinically relevant endophenotypes of MS, with genetic cluster 2 patients having a more severe phenotype and higher risk of disability. Moreover, genetic stratification is able to predict response to DMTs and could potentially be used for precision medicine in MS management.

## Introduction

Multiple sclerosis (MS) is a neuro-inflammatory and degenerative disorder with heterogenous disease outcomes. More than 15 disease modifying treatments (DMT) are approved, with a range of efficacy and safety. Accurate risk prediction of inflammation and development of sustained disability is therefore important for personalised disease management but remains difficult to determine at disease onset ^1^.

Over the last two decades, our understanding of the genetics of MS has improved. 233 single nucleotide variants (SNVs) are associated with MS susceptibility ^2^. In addition, one genome wide significant and several suggestive loci increase risk of a more severe disease course ^3,4^. However, genetic studies assessing disease outcomes have focused on single variants, with conflicting results ^5,6^.

In other disease areas ^7,8^ clustering of patients based on genetic variants has increased accuracy of predicting outcomes, although this approach has never been employed in MS. We integrated results of three pivotal susceptibility and progression genome wide association studies (GWAS) ^2–4^ and applied unsupervised hierarchical genetic clustering. We associated the resultant patient clusters with clinical outcomes in a cohort of people with MS (pwMS, n=1455) for whom detailed longitudinal phenotypic data was available. Results were then replicated and expanded within the Netherlands Brain Bank– MS cohort (NBB-MS, n=272, Fig. 1).

**Figure 1:**
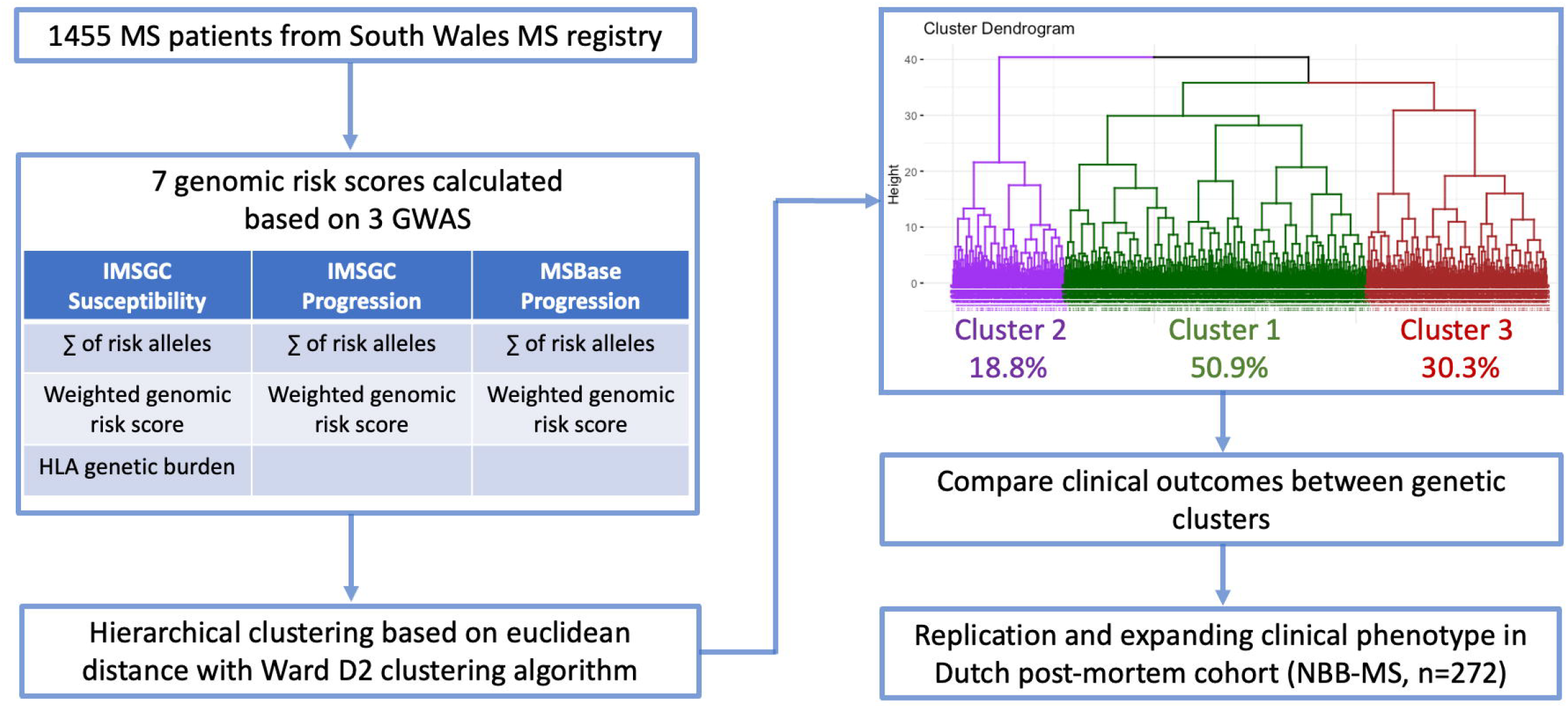
overview of the study. 1,455 MS patients were included and we calculated 7 genomic risk scores based on 3 pivotal MS GWAS. Unsupervised hierarchical clustering (Ward D2) was applied on the genomic risk scores. 3 Genomic clusters of pwMS were identified and patients within those clusters were compared with regards to relevant clinical outcomes

## Materials and methods

### Participants

1455 patients were recruited from the South Wales MS Registry established in 1985. Consecutive patients undergo annual assessment and during each visit, demographic and clinical data (including DMT, MRI and Expanded Disability Severity Score, EDSS) are collected by trained physicians recorded in a standardised web-based form. Inclusion was limited to Caucasian pwMS with an established diagnosis according to 2017 McDonald Criteria ^9^. This study was approved by the Wales Research Ethics Committee.

The replication cohort consisted of Caucasian donors with post-mortem confirmed MS (n=272) from the Netherlands Brain Bank (NBB). The study was approved by the Free University Medical Center Medical Ethics Committee. Clinical, neuropathological, and genetic information from donors were obtained, cleaned, and linked to ontology structures, within the Netherlands Neurogenetics Database. All participants provided written informed consent ^10^.

### Genotyping

Welsh cohort genotyping methodology has been described elsewhere ^5^. Briefly, DNA from EDTA-containing tubes (BD) was stored at −80°C. Genotyping was performed on either Infinium CoreExome-24 v2 or v3 chip or Immunochip (all Illumina) according to manufacturers’ instructions. Stringent quality criteria were applied, and genotypic data was imputed.

For the NBB-MS cohort, genotyping was performed on blood or brain tissue using Infinium Global Screening Array (GSA) v.3.0 according to manufacturers’ instructions. Related and non-European donors were excluded after applying quality control and imputation as previously described ^11^.

### Assessment of risk of collider bias

We combined all genomic risk scores and assessed if the different genomic risk scores are independent of each other to prevent collider bias (a biased association between a genetic variant associated with susceptibility and having an effect on severity) conditioned on a common effect (in this case developing MS). Summary statistics for both the IMSGC susceptibility and progression GWAS and the MSBase progression GWAS were obtained. Miami plots were constructed and the significant and suggestive SNVs for the progression GWAS were plotted and visually inspected if a variant was not significant in the other GWAS ^12^. To improve visibility of all non-major histocompatibility SNVs, the axis level of significance (y-axis) for susceptibility GWAS was capped at 5*10^−25^. In addition, we applied Slope Hunter adjustment for collider bias ^13^.

### Genomic risk scores and unsupervised hierarchical clustering

All available SNVs (Supplementary table 1) were used from the MS susceptibility GWAS ^2^ (significant findings only). For the progression GWAS ^3,4^ both significant and suggestive loci were used due to limited power. The number of risk alleles per patient and weighted genomic risk score were calculated. Total weighted genomic risk score (wGRS, the total genetic risk for MS per individual patient) was calculated using 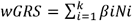, where N is the number of risk alleles and β the logit of the OR of SNV i ^14^. Human leukocyte antigen (HLA) genomic burden score was calculated based on the MS susceptibility GWAS (2). We choose to apply unsupervised clustering rather than supervised artificial intelligence models (for example random forest modelling), because the latter requires a well-defined outcome, which is challenging in MS. We compared different unsupervised hierarchical clustering algorithms using agglomerative coefficient and selected the algorithm with the highest coefficient (euclidean distance with Ward D2 clustering algorithm) according to a pre-specified analysis plan. Only the results of this clustering algorithm were taken forward to compare clinical characteristics and outcomes of pwMS based on patient’s assigned genomic cluster (Fig. 1).

### Relapse and EDSS

Annualised relapse (ARR) rate was calculated by dividing the total number of relapses by follow-up in years for each patient. Individuals with primary progressive disease were excluded from analysis. EDSS obtained during relapse were excluded from analysis (to prevent overestimation of disability due to the possibility of a temporary worsening in neurological function).

### Imaging data

MRIs were collected as part of routine clinical care. Number of T2 lesions were collected longitudinally in a standardised web-based form. If this was not described, trained physicians assessed and compared scans. Baseline number of T2 lesions were dichotomised into < 9 or ≥9. We calculated annual increase in T2 lesion burden for patients who underwent serial MR imaging at least every 2 years.

### DMT

Patients were categorised into groups who were commenced on moderate efficacy (all interferons, teriflunomide, fumarates and glatiramer acetate), high efficacy (cladribine, S1P inhibitors, monoclonal antibodies), or patients switching from moderate to high efficacy DMT or never exposed to DMTs ^15^.

### Multi-level modelling

Covariates included sex, age of onset, and genotype clusters. Time since onset (years) was used as a time metric, DMTs are time-varying covariates as they are administered at different time points. Factional polynomials were utilised to account for non-linear disease progression in the model. To avoid modelling EDSS during relapse, scores within 6-months post-relapse were excluded. To reduce autocorrelation caused by short-interval observations or prolonged unchanged periods, we summarised observations within quarter-year intervals using the median score before model fitting. Variation within individuals may depend on time, allowing nonconstant residual variance. We accounted for this using a complex level 1 variance model, modelling level 1 residuals as a function of time, previously described ^16^.

### Survival analysis

Primary endpoint was time to develop sustained disability using EDSS milestones (4, 6 and 8) or time to develop secondary progressive MS (SPMS, determined based on an increase in EDSS as previously described, in relapsing onset patients only ^5^), adjusted for sex, age at onset and DMT use. Patients were assigned to a genomic cluster which was used as an independent predictor. We constructed multivariate Cox Proportional Hazard models with genomic clusters and well-known clinical risk factors for disease progression (cerebellar disease onset, number of relapses in first 5 years, and optic neuritis as protective factor) (1). We computed three hazard ratios (HR) to assess if genomic clustering adds predictive value for the risk to develop EDSS milestones, the reference groups were resp. (i) patient in genetic cluster 1 presenting without a cerebellar syndrome, (ii) patients presenting with an optic neuritis and genomic cluster 1 and (iii) patients in genomic cluster 1 with ≤2 relapses in the first five years after onset of symptoms.

### Symptomatology, comorbidities and EDSS in the NBB-MS cohort

Survival analysis for time to EDSS 6 and other clinical outcomes were performed as in the Welsh cohort. Differences in disease progression might also manifest as differences in experienced symptomatology. For the NBB-MS cohort, neuro(-psychiatric) signs and symptoms were scored in medical record summaries of brain donors through natural language processing as described previously ^10^. We compared the number of observations of a given sign or symptom between the genetic clusters, assessed difference in their temporal distribution (both Kruskal Wallis test), and performed a survival analysis for each symptom (log-rank test). Comorbidities are potential confounders of disease progression. For the NBB-MS cohort, comorbidity information was available. Comorbidities were grouped into higher-order categories of the Human Disease Ontology. For each cluster, the proportion of donors with at least one member of each higher-order category was calculated and compared (χ^2^ test).

### Statistical analysis

Comparison of baseline characteristics was undertaken using non-parametric analysis (Kruskal Wallis test). Correlation between the genomic scores was calculated using Spearman’s rho. All statistical analyses were performed with R Studio (Supplementary materials). We applied False Discovery Rate (FDR) correction for multiple testing if necessary and adjusted p-value <0.05 were considered statistically significant.

## Results

### Individual genomic risk scores are not associated with time to sustained disability

We assessed whether different genomic risk scores for MS susceptibility and severity were predictive for development of sustained disability. We did not observe a consistent association between a genomic risk score and the HR to develop EDSS 4, 6, and 8 or SPMS (Supplementary table 2-5), validating another study using MS susceptibility SNPs to predict disease severity ^17^, indicating that single genomic risk scores are unable to predict MS disease course.

### No evidence of collider bias between susceptibility and progression GWAS

We constructed Miami plots for several MS genetic risk scores and found no evidence for shared genetic effects (Fig. 2A-B). After applying a Slope Hunter adjustment for potential collider bias, we did not observe a deviation of the beta of the progression GWAS (Fig.2C), indicating that it is unlikely that collider bias could affect the results of this study. In addition, no significant correlations were found between the genetic risk scores (Fig. 2D).

**Figure 2:**
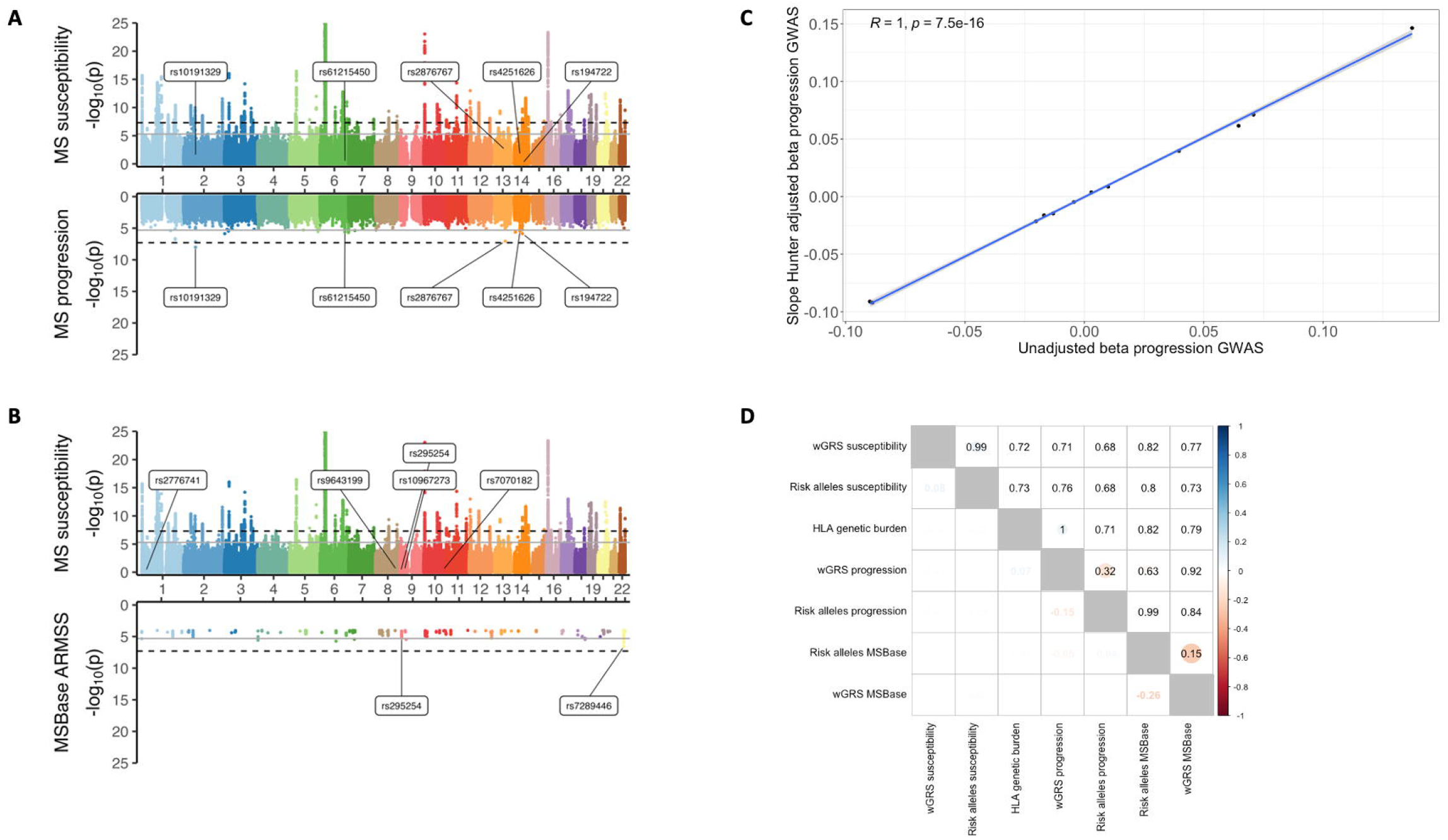
no evidence for collider bias. Miami plots to assess that genetic loci are independent of each other. **A)** IMSGC susceptibility GWAS vs IMSGC progression GWAS, **B)** IMSGC susceptibility GWAS vs MSBase GWAS **C)** Slope Hunter adjustment betas showed similar betas compared to unadjusted beta obtained from the IMSGC progression GWAS, indicating that collider bias is not affected our results. **D)** Correlation plot between different genomic risk scores, with correlation Spearman’s correlation coefficient in the lower triangle and associate p-values in the upper triangle.

### Unsupervised hierarchical clustering on genomic risk scores identified three clusters of patients with similar genomic risk profiles

We applied unsupervised hierarchical clustering solely based on genomic data in the Welsh cohort. Using this algorithm, we obtained a Ward agglomerative coefficient of 0.97, indicating that patients are well-clustered (Fig. 1). No significant differences in the genomic risk scores were found between the Welsh and NBB-MS cohort (all p>0.05), indicating robust and generalisable results between the two cohorts. Next, we validated that the genomic risk scores were significantly different between different clusters and all scores independently contribute to the clustering approach (Figure 3A-H), all p-values <7.4*10^−8^). Cluster 3 has the strongest genetic risk for MS susceptibility (Fig. 3D) ^2^, with the largest number of risk alleles. Cluster 1 has the highest HLA burden (Fig. 3G) ^2^, cluster 2 has the most risk alleles from the MSBase progression GWAS (Fig. 3C) ^4^.

**Figure 3:**
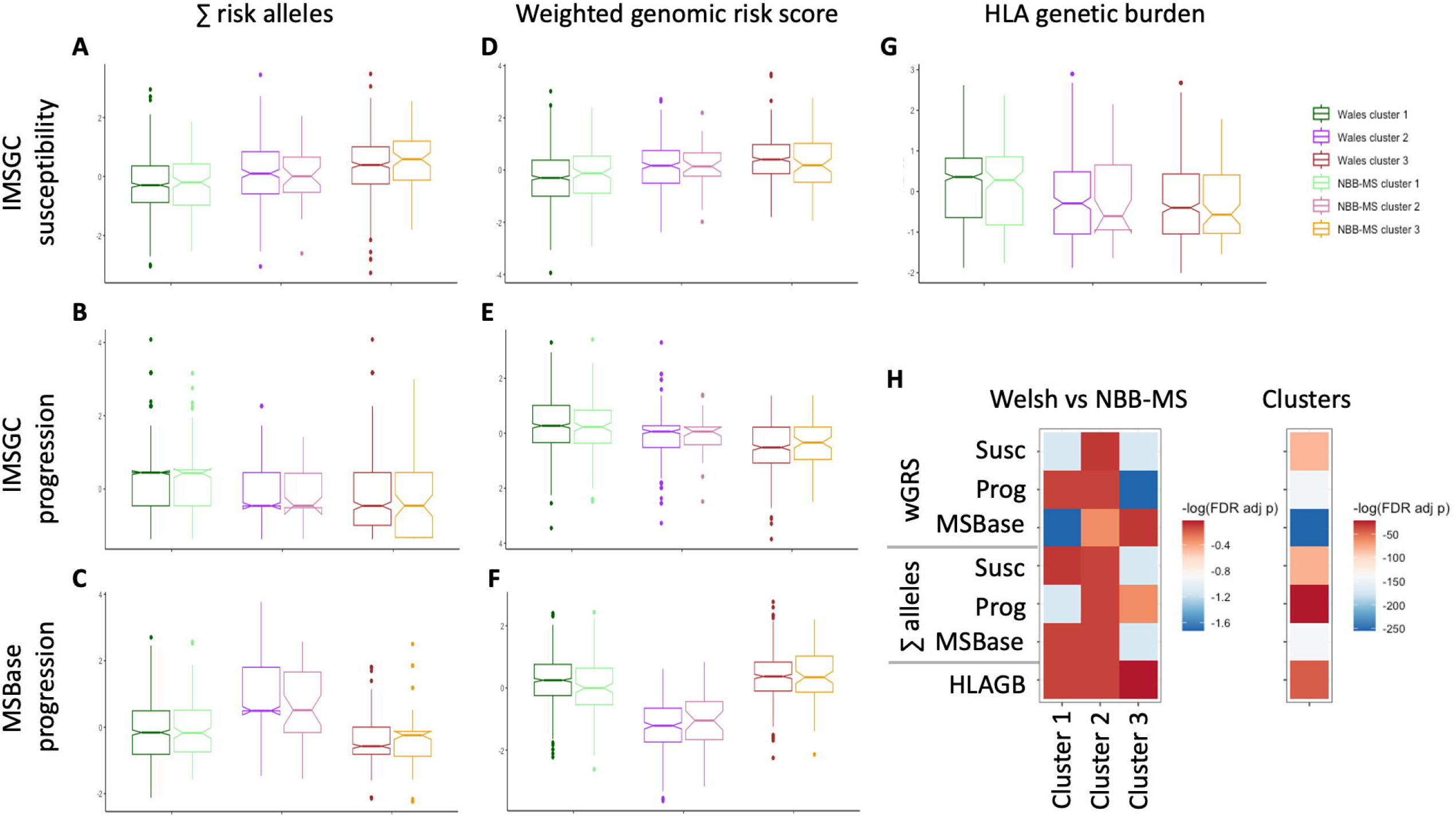
genomic clustering differentiates genetic risk scores and shows comparable patterns between Welsh and NBB-MS cohort. Comparison of genomic risk scores between the genetic clusters, in every figure left boxplot represent Welsh MS patients and right boxplot Dutch MS patients. **A)** sum of risk alleles of susceptibility variants, B) sum of number of risk alleles of severity variants of IMSGC GWAS, **C)** sum of risk alleles of MSBase GWAS, D) weighted genomic risk score for susceptibility variants, **E)** weighted genomic risk scores for IMSGC progression GWAS, **F)** weighted genomic risk scores for MSBase GWAS and **G)** Human leukocyte antigen (HLA) genetic burden score. **H)** Heatmap of FDR adjusted p-values, left panel comparing within a genetic cluster the genomic scores between the Welsh and NBB-MS cohort and ANOVA results comparing the genomic risk scores between the three genetic clusters for the pooled Welsh and NBB-MS cohort.

### Severity of MS at disease onset is similar between genomic clusters

No significant differences in age at onset, gender or number of T2 brain lesions on MRI were identified between clusters, indicating that severity of disease at onset is independent of genetic background (Table 1-2) and reflects established difficulties of predicting long-term disease outcomes at baseline.

**Table 1:** Characteristics at disease onset similar between genomic clusters.

|  | South Wales cohort (n=1455) |  |  | p-value | Dutch cohort (n=272) |  |  | p-value |
| --- | --- | --- | --- | --- | --- | --- | --- | --- |
|  | Cluster 1<br>(n=750) | Cluster 2<br>(n=277) | Cluster 3<br>(n=446) |  | Cluster 1<br>(n=145) | Cluster 2<br>(n=38) | Cluster 3<br>(n=84) |  |
| Age at onset | 33.0<br>(10.9) | 33.5<br>(10.8) | 33.1<br>(10.6) | 0.76 | 32 (15.5) | 32 (20) | 32.5<br>(14) | 0.80 |
| Percentage female | 70.6% | 71.1% | 67.9% | 0.53 | 67.6% | 68.4% | 63.1% | 0.37 |
| Months between first and second relapse<sup>1</sup><br>(median, IQR) | 22 (60) | 23.3<br>(59.9) | 61.6<br>(47) | 0.25 | - | - | - | - |
| Oligoclonal bands positive/number tested<br>(positive %) | 367/449<br>(81.7%) | 142/167<br>(85.0%) | 233/282<br>(82.6%) | 0.63 | - | - | - | - |
| Percentage PPMS | 10.9% | 10.3% | 9.2% | 0.67 | 22.1% | 31.6% | 20.2% | 0.37 |
| First EDSS after onset (median, IQR) | 3.5 (3) | 3.5 (3.5) | 3.5 (3.5) | 0.97 | - | - | - | - |
| Disease modifying treatments<br>(number of patients) |  |  |  |  | - | - | - | - |
| Never | 546 | 208 | 323 |  |  |  |  |  |
| Only moderate efficacy | 126 | 47 | 80 |  |  |  |  |  |
| Only high efficacy | 35 | 10 | 20 | 0.95 |  |  |  |  |
| Switch between moderate and high efficacy | 43 | 12 | 23 |  |  |  |  |  |
| Age at death <sup>2</sup><br>(mean, sd) | n=76<br>63.3<br>(12.4) | n=35<br>59.5<br>(13.0) | n=55<br>63.0<br>(9.9) | 0.38 | 64.5(12.9) | 62.1<br>(13.9) | 61.8<br>(11.2) | 0.12 |

<sup>1</sup> Relapsing onset patients only, Cohen's d comparing different clusters negligible (max 0.17); <sup>2</sup> ANOVA

**Table 2:**
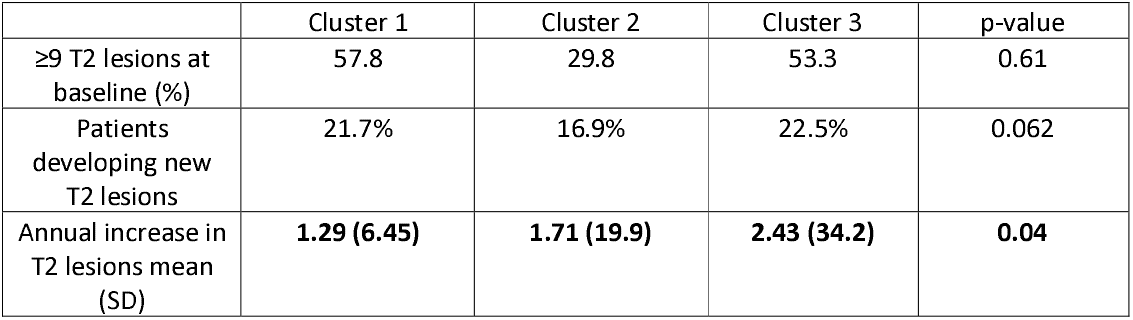
MS patients in genomic cluster 2 and 3 have a significantly higher annual increase in T2 lesion load on serial MR imaging.

|  | Cluster 1 | Cluster 2 | Cluster 3 | p-value |
| --- | --- | --- | --- | --- |
| ≥9 T2 lesions at baseline (%) | 57.8 | 29.8 | 53.3 | 0.61 |
| Patients developing new T2 lesions | 21.7% | 16.9% | 22.5% | 0.062 |
| Annual increase in T2 lesions mean (SD) | **1.29 (6.45)** | **1.71 (19.9)** | **2.43 (34.2)** | **0.04** |

### Patients in genomic cluster 2 have significantly increased risk of developing fixed disability

We compared long-term disease outcomes and found no significant differences in time to impaired ambulation (EDSS 4) between different genetic clusters (Supplementary figure 1). Interestingly, patients in cluster 2 and 3 had a significantly shorter time to requiring a walking aid (EDSS 6, 5.7 and 4.9 years earlier respectively, Fig 4A) and time to wheelchair dependency (EDSS 8, 2.8 and 16.2 years earlier respectively Fig. 4B) compared to cluster 1. In relapse onset patients, time to develop SPMS was significantly shorter for patients in genomic cluster 2 and 3 (3.9 and 5.1 years respectively Fig. 4C). Patients in genomic cluster 2 and 3 also had a significantly higher risk of developing EDSS 6 (HR 1.32, p=0.009 and 1.46, p=4*10^−5^, Supplementary table 3), EDSS 8 (HR 1.45, p=0.038 and 1.49, p=0.0087, Supplementary table 4) and SPMS (HR 1.27, p=0.030 and 1.30, p=0.0063, Fig. 4E and Supplementary table 5). We then assessed if genomic clustering influenced the number of relapses and whether this could contribute to development of sustained disability. No significant differences were found between genomic clusters in annualised relapse rate in RRMS (Cohen’s d max 0.078, p=0.23, data not shown), indicating that the genetic variants are likely related to accelerated development of sustained disability independent of relapse activity. Next, we compared EDSS trajectories using a multi-level model to compare the annual increase in EDSS. We included 8,941 non-relapse associated EDSS scores from 1,029 relapsing pwMS. Nine years post-onset of MS, EDSS trajectories started to diverge, patients in genomic cluster 2 had an 0.38 (95% CI 0.00075-0.75, Fig 4F and Supplementary table 6) increase in EDSS compared with cluster 1. Finally, we replicated time to EDSS 6 in the independent NBB-MS cohort; patients in genomic clusters 2 and 3 had an accelerated time to development of sustained disability (p=0.044, Fig. 4D) independent of comorbidities (Fig. 4G), validating that genomic clusters 2 and 3 are associated with a more severe disease course.

**Figure 4:**
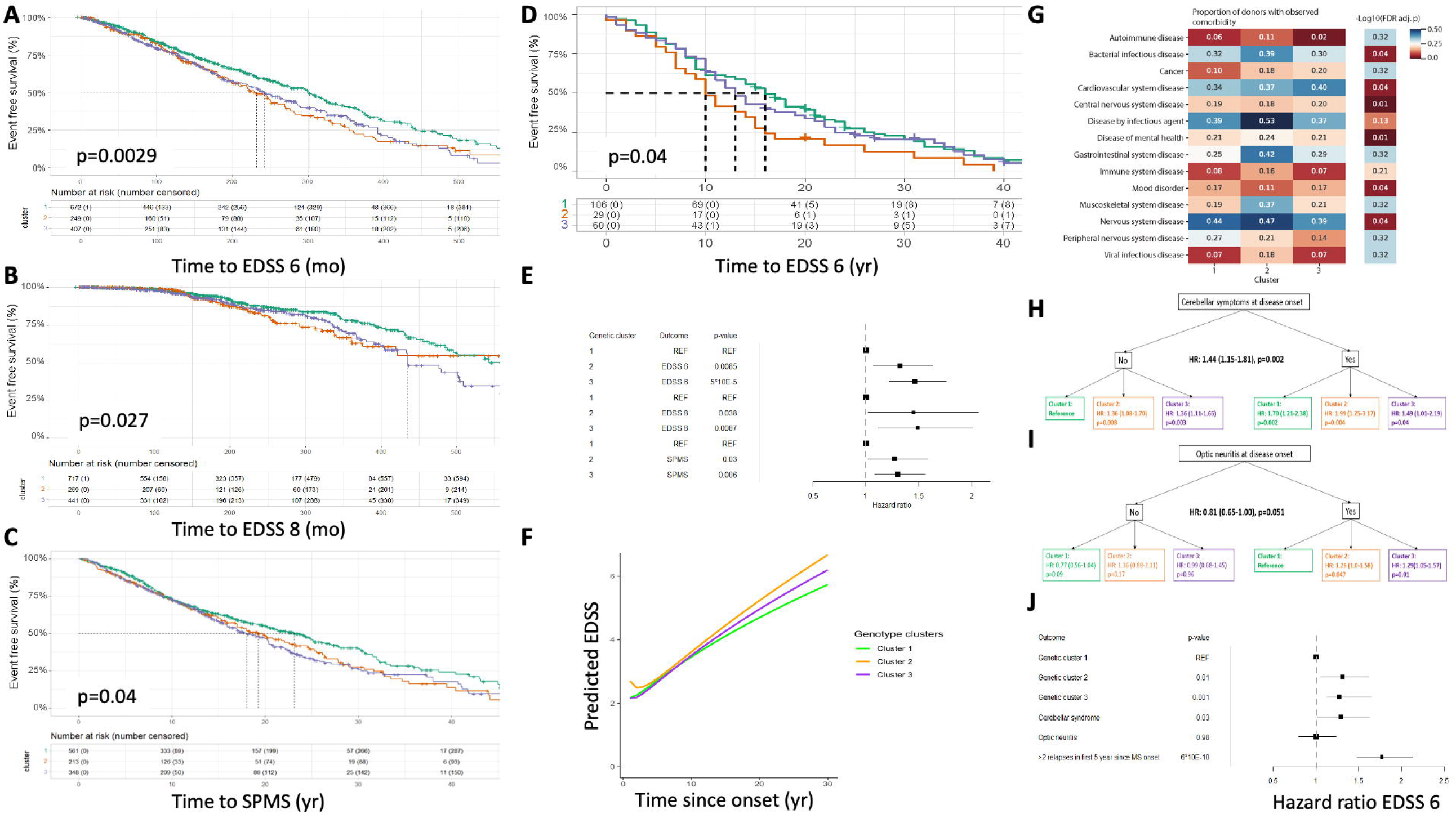
patients in genomic cluster 2 have significantly worse disease outcomes. Kaplan-Meier survival curves for time to **A)** EDSS 6, **B)** EDSS 8 and **C)** SPMS in the Welsh cohort. **D)** Kaplan Meier curve for time to EDSS 6 in the NBB-MS cohort **E)** Hazard ratios to develop several disability milestones, adjusted for gender, age at onset and use of DMT. **F)** Multi-level modelling to compare the changes in EDSS over time between the different genetic clusters. **G)** Heatmap for comorbidities comparing the genetic clusters in the Dutch cohort. Hazard ratios of genetic clusters compared to well-validated clinical risk factors; **H)** cerebellar syndrome or **I)** optic neuritis at disease onset. **J)** Multivariate Cox Proportional Hazard model for hazard ratio to develop EDSS 6, adjusted for sex, age at onset and use of DMT.

### Genomic clustering is an independent predictor of disease progression compared to well-validated clinical risk factors

We assessed performance of genomic clustering to predict risk of disease progression compared to well-validated clinical predictors ^1^. We found that patients presenting with a cerebellar syndrome at disease onset overall have a HR of 1.44 of developing EDSS 6. Interestingly, patients without cerebellar syndrome at onset (favourable outcome based on current risk prediction) within genomic cluster 2 and 3 had a similar risk of developing EDSS 6 as patients presenting with cerebellar signs (Fig. 4H). We found a similar pattern that genomic clustering improves risk prediction for patients with two or less relapses within the first 5 years after onset (Supplementary figure 2). Patients with optic neuritis at onset overall had a lower risk of developing EDSS 6 (HR 0.81). However, optic neuritis patients within genetic cluster 2 and 3 had a significantly increased risk of developing EDSS 6 compared to cluster 1 (HR 1.3, Fig. 4I). Therefore, genetic testing at onset provides additional predictive information for the risk of sustained disability (Fig. 4J).

### Genomic cluster 1 patients have significantly lower annualised increase in T2 lesions on MRI

539 pwMS had at least bi-annual brain MR imaging. At baseline, no differences were found in the percentage of patients having 9 or more lesions between the different genomic clusters (p=0.61). During follow-up, no apparent differences between the genetic clusters were found for the chance of new lesions (p=0.062). However, comparing annualised increase in T2 lesions in patients having active MRI revealed that patients in genomic cluster 2 and 3 had a significantly higher number of new lesions compared to cluster 1 patients (1.71 and 2.43 new T2 lesions per annum, p=0.04, Table 2).

### Patients in genomic cluster 2 have best response to treatment

Time to develop disability milestones may be delayed by DMTs. Therefore, we assessed effect of DMT within our genetic clusters comparing patients who had never been treated versus patients ever receiving a DMT. Genetic cluster 2 patients who received a DMT had a significantly longer time to develop EDSS 6 compared to untreated patients (Fig. 5A, p=0.004) and similar effects were found for time to EDSS 8 (p=0.005, Fig. 5B) with a trend towards significance for SPMS (p=0.06, Fig. 3C). In contrast, we only observed an effect of an MS DMT in cluster 1 patients for time to EDSS 8 (Fig. 5B) and no effect of a DMT in cluster 3 patients (Fig. 5C). These effects were replicated in the multi-level model (Supplementary table 6). We did not observe a difference in time to EDSS 4 between treated and untreated patients between the different genomic clusters (Supplementary figure 3). Genomic clustering may help identify good responders to DMTs.

**Figure 5:**
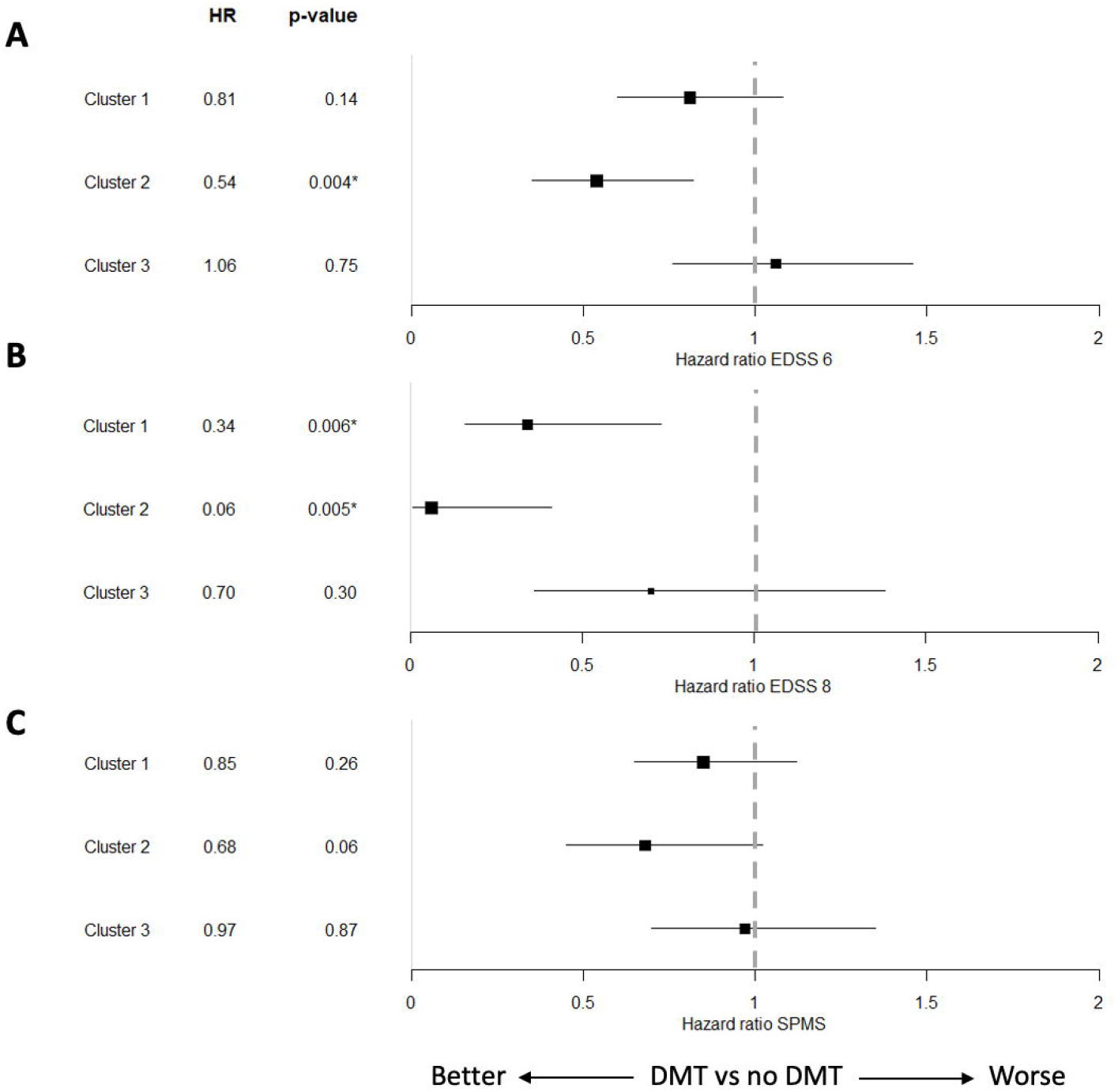
patients in genetic cluster 2 have the best response to disease modifying treatments. Within each genomic cluster, we compared time to disability milestones between patients who ever received a DMT versus patients who have never been treated, and calculated hazard ratios for time to **A)** EDSS 6, **B)** EDSS 8 and **C)** SPMS

### Patients in genomic cluster 1 have a more favourable disease phenotype

Finally, we assessed whether genomic clustering was associated with a more severe disease phenotype in the NBB-MS cohort, beyond symptoms (easily) captured with EDSS assessment. Genomic cluster 1 patients had a more favourable disease course on a broad range of symptoms, including cognition, bladder involvement and fatigue (Fig. 6A) ^10^. Patients in cluster 2 were younger when they developed motor and non-motor symptoms (Fig. 6B-D) and did so more frequently (Fig. 6E-F), indicating that genetics is associated with disease phenotype.

**Figure 6:**
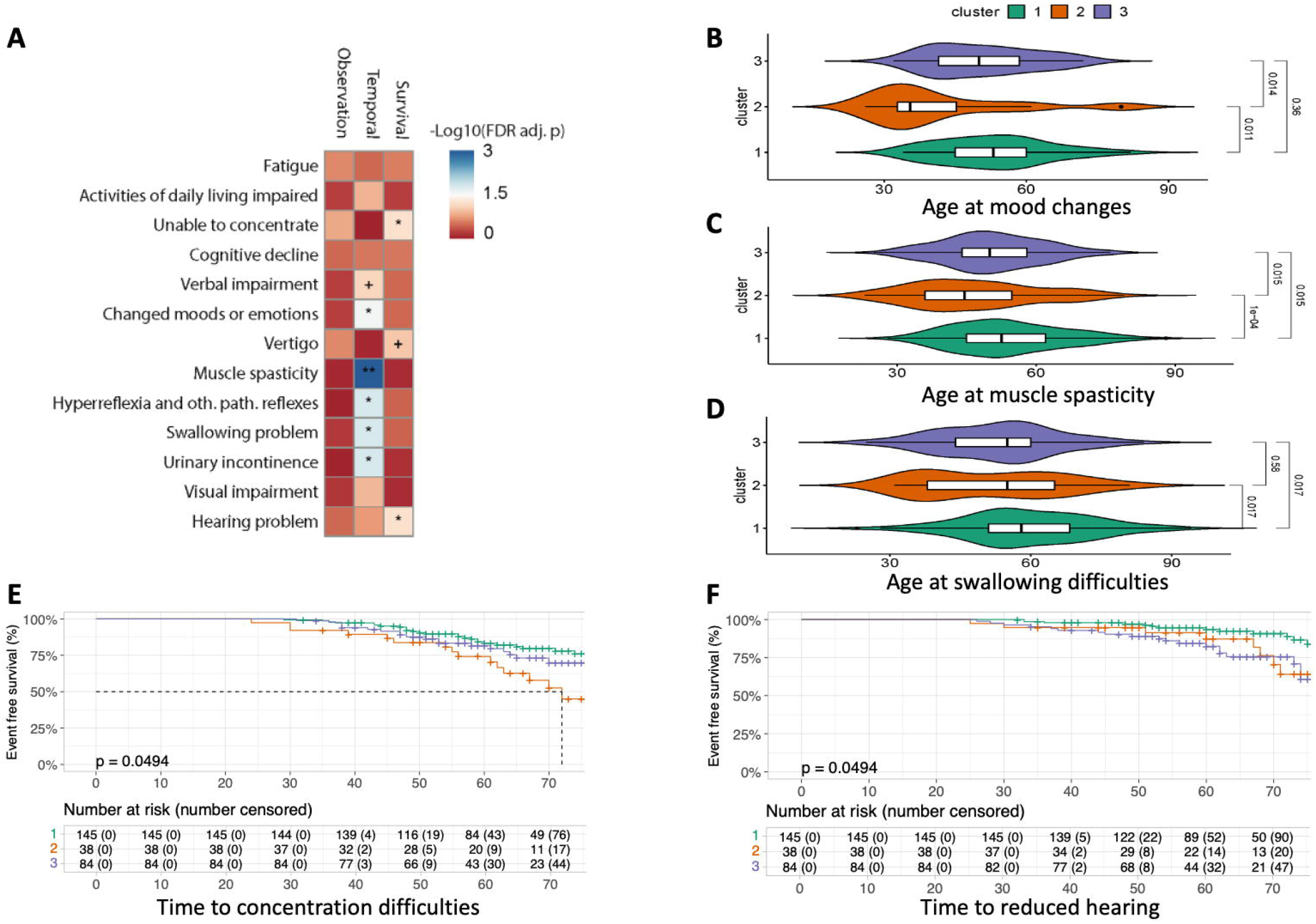
genetic cluster 1 patients have the most favourable disease course. Natural language processing was applied to medical records of the Dutch cohort. **A)** We compared the number of observations of a specific symptom (first column of the heatmap), the age occurrence of the symptoms (second column) and differences in event free survival (third column), all p-values are FDR corrected for multiple testing. Differences in age at onset were compared between the three different clusters for **B)** changes in mood, **C)** development of muscle spasticity, **D)** and swallowing difficulties. Survival analysis for time **E)** difficulties with concentration and **F)** reduced hearing.

## Discussion

Based on unsupervised genomic clustering using the results of three pivotal GWAS in MS, we have identified three patient clusters with similar baseline characteristics but different disease trajectories, in two independent cohorts. Analysis of longitudinal follow-up data revealed that patients in cluster 2 and 3 had a higher annual increase in T2 lesion load on serial MR imaging and a higher risk of developing early fixed disability during their disease course, independent of relapse activity. Patients in cluster 2 showed the best response to DMT. We confirmed that patients in genomic cluster 2 have a worse phenotype compared to cluster 1 in a second independent cohort of patients. While one could consider this second cohort small when performing unbiased genetic analysis, we could replicate our findings with regards to time to EDSS 6, because of the large effect sizes we observed when applying genomic clustering. In addition, to prevent a self-fulling prophesy, we could not use patients who have been included in the MS progression GWASs ^2–4^. These data suggest that genomic clustering provides a valuable tool to detect endophenotypes of MS and to increase accuracy of prognostication and inform management decisions in MS.

To our knowledge, this is the first study applying unsupervised clustering analysis integrating all currently available genetic information into one prediction model for MS. In other disease areas, genomic clustering has been shown to identify relevant disease phenotypes. In patients with insulin resistance, genomic clustering was able to identify subgroups of people who had a higher risk of developing hypertension and coronary artery disease, whereas another cluster was associated with altered lipid metabolism ^8^. In patients with type 2 diabetes, genomic clustering was associated with a subgroup with more insulin resistance, and another cluster showed reduced insulin secretion ^7^. In addition, in a large study involving Alzheimer’s, Parkinson’s disease (PD), frontotemporal dementia and amyotrophic lateral sclerosis, genomic clustering revealed novel insights in underlying pathologies ^18^. By integrating clinical and genomic data from PD, Alzheimer’s disease risk loci were shown to be related to clinical phenotypes in PD ^19^ and also clustering on clinical data can identify subgroups of patients in primary lateral sclerosis with novel genetic mutations ^20^.

In the current study, MS patients in genomic cluster 2 and 3 had a significantly shorter time to developing sustained disability. These two patient groups also had the highest annual increase in T2 lesion load. The development of new T2 lesions on MRI is a well-known risk factor for developing sustained disability ^21–23^ and could, at least partially, be driven by genetic factors. Another interesting observation was that using newly developed multi-level modelling ^16^, within 9 years of disease onset, the rate of increase in EDSS is similar between genomic clusters. However after 10 years, particularly in cluster 2, there was accelerated disability accumulation, validating the non-linear disease trajectory of MS and underlining the importance of genetics in disability progression independent of relapse activity.

Our study has limitations. We observed differences in response to treatment, but we were unable to stratify this effect to specific DMTs because of small patient numbers. Larger studies, preferably nested within a clinical trial, will need to address this further. Secondly, the inter-rater variability of EDSS is relatively high ^24^ and could lead to some ascertainment bias. Collection of EDSS measurements from 1985-2022, used different physicians. However, considering the high number of EDSS per patients, we do not expect any related systemic bias. In addition, accumulation of T2 lesion load and non-EDSS outcomes all showed similar patterns with patients in cluster 2 having a more severe disease course compared to cluster 1. Thirdly, because we included a historical cohort, not all pwMS underwent regular MR imaging. The imaging-based outcomes could therefore only be assessed in a subgroup of patients over-represented by those receiving a DMT. However, this would be more likely to lead to an underestimation of annualised increase in T2 lesion load, since untreated patients would be more likely to develop new lesions compared to treated patients. Finally, patients in genetic cluster 2 had a significantly shorter time to EDSS milestones, but the best response to treatment. This could reflect some degree of residual confounding by indication or collider bias and the effects of DMT on progression are not causal effects as we have not adjusted for all confounding. In addition, a part of the included patients were diagnosed before the licensing of DMT and therefore the lag time between onset of the disease and the start of their DMT could contribute to a diminished response to treatment (e.g. because of the delayed commencement of their treatment, those patients had an increased risk of disease progression due to a higher annualised relapse rate ^25^). Future research could explore methods that account for time-varying treatments and confounders (such as marginal structural models) ^26^ to robustly assess whether these genetic clusters can modify the effect of DMTs.

In summary, we have identified three genetic clusters that improve the accuracy of predicting long-term disease outcomes and could potentially serve as a prognostic factor to guide precision medicine in MS. Further studies are necessary to understand the underlying biology (which genetic loci are driving the effect and which pathways are affected) and to develop more targeted therapeutic approaches.

## Data Availability

All data produced in the present study are available upon reasonable request to the authors and upon completion of a signed data transfer agreement.

## Acknowledgement

We are grateful to Prof. Davey Smith, Karen Ho BSc and Dr. Tom Clark (University of Bristol, UK) for assistance with the genotyping and all people with MS who have participated in this study. We would like to acknowledge the staff from the Netherlands Brain Bank, as well as the brain donors and their families, for their invaluable contributions to science.

## Authors contribution

KLK, ECT, NPR contributed to the conception and design of the study; KLK, NJM, EU, SL, RWT, KEH, PH, JWJB, ECT, IRH, NPR contributed to the acquisition and analysis of data; KLK, NJM, EU, SL, RWT, KEH, PH, JWJB, ECT, IRH, NPR contributed to drafting the text or preparing the figures.

## Potential Conflicts of Interest

None of the authors report a conflict of interest.

## Notes

### Competing Interest Statement

The authors have declared no competing interest.

### Funding Statement

This study did not receive any funding

### Author Declarations

1455 patients were recruited from the South Wales MS Registry established in 1985. This study was approved by the Wales Research Ethics Committee. The replication cohort consited of Caucasian donors with post-mortem confi rmed MS (n=272) from the Netherlands Brain Bank (NBB). The study was approved by the Free University Medical Center Medical Ethics Committee.

