## Supplementary materials for "Genetic subtypes predict multiple sclerosis severity and response to treatment"

**Supplementary data**

Karim L Kreft ^1,2^, Nienke J. Mekkes ^3,4^, Emeka Uzochukwu ^2^, Sam Loveless ^2^, Ray Wynford-Thomas ^2^, Katharine E. Harding ^5^, Mark Wardle ^1^, Peter Holmans ^2^, J. William L. Brown ^6^, Michael Lawton ^7^, Emma C Tallantyre ^1,2^, Inge R. Holtman ^3,4^, Neil P Robertson ^1,2^

Affiliations

^1^ Department of Neurology, University Hospital of Wales, Cardiff, UK.

^2^ Division of Psychological Medicine and Clinical Neuroscience, Cardiff University, Cardiff, UK.

^3^ Department of Biomedical Sciences, Section Molecular Neurobiology, University of Groningen, University Medical Center Groningen, Groningen, The Netherlands

^4^ Machine Learning Lab, Data Science Center in Health, University of Groningen, University Medical Center Groningen, Groningen, The Netherlands.

^5^ Department of Neurology, Royal Gwent Hospital, Newport, United Kingdom

^6^ Department of Clinical Neurosciences, University of Cambridge, Cambridge, United Kingdom

^7^ Bristol Population Health Science Institute, Bristol Medical School, University of Bristol, Bristol, United Kingdom

Corresponding authors:

Karim L. Kreft,

Neil P. Robertson,

*Additional materials and methods*

Rstudio version 2023.06.1 with base R version 4.2.2, except multi-level modelling, which was done in version 4.1.1)

Packages used

- data.table (1.14.6)
- tidyverse (1.3.2, including ggplot2 to create the graphs)
- readxl (1.4.2)
- survival (3.4.0)
- survminer (0.4.9)
- luster (2.1.4)
- miamiplot (1.1.0)
- corrplot (0.92)
- janitor (2.2.0)
- rstatix (0.7.2)
- SlopeHunter (1.1.0)
- R2MLwiN (0.8.8)
- lme4 (1.1.35.1).

*Supplementary table 1: Available SNV after QC and imputation to calculate genomic risk scores for IMSGC susceptibility and progression GWAS, and MSBase GWAS*

| rs12588969 | rs13136820 | rs34695601 | rs6837324 | rs7454108 |
| --- | --- | --- | --- | --- |
| rs6427540 | rs1323292 | rs34947566 | rs6928313 | rs9266629 |
| rs13385171 | rs13327021 | rs354033 | rs6952809 | rs9277626 |
| rs10063294 | rs13551 | rs35486093 | rs6990534 | rs10191329 |
| rs1014486 | rs1365120 | rs35540610 | rs6999228 | rs61215450 |
| rs10171296 | rs137955 | rs35703946 | rs701006 | rs2876767 |
| rs10191360 | rs1399180 | rs36090551 | rs7014582 | rs4251626 |
| rs10230723 | rs140522 | rs3737798 | rs706015 | rs194722 |
| rs10245867 | rs1415069 | rs405343 | rs719219 | rs7289446 |
| rs1026916 | rs1465697 | rs4262739 | rs719316 | rs10967273 |
| rs10271373 | rs1534422 | rs4325907 | rs7222450 | rs698805 |
| rs1077667 | rs16902700 | rs438613 | rs7260482 | rs295254 |
| rs10801908 | rs17051321 | rs4409785 | rs72922276 | rs9643199 |
| rs1087056 | rs1738074 | rs4728142 | rs72928038 | rs2776741 |
| rs10914539 | rs17741873 | rs4743150 | rs72989863 | rs7070182 |
| rs10936602 | rs17780048 | rs4772201 | rs73414214 | rs12614091 |
| rs10951042 | rs17797448 | rs4796224 | rs735542 | rs12622670 |
| rs11079784 | rs1800693 | rs4808760 | rs767455 | rs12708716 |
| rs11083862 | rs1801133 | rs4812772 | rs7731626 | rs12722559 |
| rs11125803 | rs2027982 | rs483180 | rs7975763 | rs13066789 |
| rs1112718 | rs2084007 | rs4896153 | rs7977720 | rs2992 |
| rs11161550 | rs2150879 | rs4939490 | rs802730 | rs3007421 |
| rs11231749 | rs2248137 | rs4940730 | rs883871 | rs3184504 |
| rs11256593 | rs2255214 | rs531612 | rs9282641 | rs32658 |
| rs1151625 | rs2269434 | rs57116599 | rs9292776 | rs34681760 |
| rs11578655 | rs2286974 | rs5756405 | rs9308424 | rs6670198 |
| rs11749040 | rs2289746 | rs58166386 | rs9568402 | rs6672420 |
| rs1177228 | rs2317231 | rs58394161 | rs9591325 | rs67111717 |
| rs11809700 | rs2327586 | rs585039 | rs9610458 | rs6738544 |
| rs11852059 | rs2331964 | rs59655222 | rs962052 | rs6789653 |
| rs11919880 | rs244656 | rs6032662 | rs9808753 | rs2523500 |
| rs12132283 | rs2469434 | rs60600003 | rs983494 | rs2844482 |
| rs12133753 | rs249677 | rs6072343 | rs9843355 | rs3093982 |
| rs12147246 | rs2546890 | rs61708525 | rs9863496 | rs3135024 |
| rs12206238 | rs2585447 | rs61863928 | rs9878602 | rs3135388 |
| rs12211604 | rs2590438 | rs61884005 | rs9900529 |  |
| rs12365699 | rs2639453 | rs62013236 | rs9909593 | ***HLA genetic burden score*** |
| rs12373588 | rs2705616 | rs62420820 | rs9937051 | ***IMSGC progression GWAS*** |
| rs12434551 | rs2726479 | rs631204 | rs9955954 | ***MSBase progression GWAS*** |
| rs12478539 | rs2836438 | rs6496663 | rs9992763 | ***The remaining SNV derived from IMSGC susceptibility GWAS*** |
| rs1250551 | rs28625973 | rs6533052 | rs11751659 |  |
| rs12609500 | rs28834106 | rs6589939 | rs2229092 |  |

*Supplementary table 2: MS genomic risk scores not associated with the development of sustained disability*

|  | EDSS 4 | EDSS 6 | EDSS 8 | SPMS |
| --- | --- | --- | --- | --- |
| **Unadjusted** | | | | |
| Σ Susceptibility risk alleles  wGRS susceptibility  HLAGB  Σ severity risk alleles (IMSGC)  wGRS severity (IMSGC)  Σ severity risk alleles (MSBase)  wGRS severity (MSBase) | 0.99(0.98-1.01) p=0.37  **1.14 (1.01-1.30) p=0.035**  0.91 (0.79-1.07) p=0.25  1.06 (0.96-1.17 p=0.27  1.20 (0.23-6.26) p=0.83  0.97 (0.91-1.05) p=0.46  0.97 (0.87-1.07) p=0.52 | 1.0 (0.99-1.01) p=0.37  1.02 (0.92-1.12) p=0.77  **0.87 (0.78-0.99) p=0.027**  1.02 (0.94-1.10) p=0.13  0.37 (0.10-1.36) p=0.66  1.0 (0.94-1.05) p=0.87  0.96 (0.89-1.04) p=0.31 | 1.01 (0.99-1.03) p=0.21  1.03 (0.87-1.21) p= 0.77  0.86 (0.70-1.05) p=0.13  1.06 (0.93-1.20) p=0.40  1.29 (0.16-10.20) p=0.81  1.07 (0.98-1.18) p=0.12  1.06 (0.94-1.20) p=0.39 | 1.0 (0.99-1.01) p=0.85  1.08 (0.97-1.19) p=0.17  0.96 (0.85-1.08) p=0.46  1.01 (0.94-1.09) p=0.77  0.67 (0.18-2.49) p=0.55  1.02 (0.96-1.08) p=0.62  1.02 (0.94-1.10) p=0.67 |
| **Adjusted *** | | | | |
| Σ Susceptibility risk alleles  wGRS susceptibility  HLAGB  Σ severity risk alleles (IMSGC)  wGRS severity (IMSGC)  Σ severity risk alleles (MSBase)  wGRS severity (MSBase) | 1.0 (0.99-1.01),   p=0.99  **1.14 (1.01-1.29) p=0.04**  0.98 (0.84-1.14)  p= 0.76  1.06 (0.96-1.17)  p=0.25  1.06 (0.19-5.82) p= 0.95  0.94 (0.87-1.01)  p=0.087  0.94 (0.84-1.04) p=0.21 | 1.0 (0.99-1.01) p=0.53  1.05 (0.95-1.15) p=0.34  0.99 (0.88-1.12) p=0.90  1.03 (0.95-1.11) p=0.49  **0.22 (0.058-0.85) p=0.028**  0.97 (0.91-1.02) p=0.22  0.97 (0.89-1.05) p=0.41 | 1.01 (0.99-1.03) p=0.27  1.04 (0.89-1.22) p=0.60  0.93 (0.76-1.14) p=0.47  1.07 (0.94-1.21) p=0.31  1.02 (0.12-8.38) p=0.99  1.07 (0.98-1.17) p=0.13  1.06 (0.93-1.21) p=0.39 | 1.0 (0.99-1.01) p=0.65  1.09 (0.99-1.21) p=0.092  1.01 (0.90-1.15) p=0.83  1.01 (0.93-1.09) p=0.85  0.56 (0.15-2.13) p=0.39  1.0 (0.94-1.06) p=0.87  1.01 (0.93-1.09) p=0.86 |

Hazard ratio (95% CI), p-value not adjusted for multiple testing

*** Sex, age at onset, disease modifying treatments

*Supplementary table 3: Unadjusted and adjusted hazard ratios for risk to develop EDSS 6*

|  | South Wales cohort | | Dutch cohort | |
| --- | --- | --- | --- | --- |
|  | Hazard ratio (95% CI) | p-value | Hazard ratio (95% CI) | p-value |
| Genomic cluster  2  3 | **1.34 (1.09-1.66)**  **1.30 (1.08-1.56)** | **0.0053**  **0.0046** | **1.61 (1.05-2.46)**  1.12 (0.80-1.60) | **0.029**  0.507 |
| Genomic cluster  2  3  Female sex | **1.37 (1.11-1.69)**  **1.29 (1.08-1.55)**  **0.79 (0.66-0.93)** | **0.0031**  **0.0055**  **0.0048** | **1.609 (1.05-2.46)**  1.123 (0.80-1.58)  1.060 (0.78-1.45) | **0.029**  0.500  0.716 |
| Genomic cluster  2  3  Female sex  Age at onset | **1.34 (1.09-1.65)**  **1.47 (1.22-1.76)**  0.92 (0.77-1.08)  **1.07 (1.06-1.08)** | **0.0062**  **4*10^-5^**  0.31  **<2*10^-16^** | 1.53 (1.00-2.35)  1.12 (0.80-1.58)  1.02 (0.75-1.40)  **1.03 (1.01-1.04)** | 0.050  0.498  0.882  **2.32 × 10-4** |
| Genomic cluster  2  3  Female sex  Age at onset  Disease modifying treatment  Only moderate efficacy  Only high efficacy  Switch moderate and high | **1.32 (1.07-1.63)**  **1.46 (1.22-1.76)**  0.93 (0.79-1.10)  **1.07 (1.06-1.08)**  0.90 (0.71-1.14)  **1.65 (1.10-2.48)**  1.19 (0.82-1.73) | **0.0085**  **4*10^-5^**  0.40  **<2*10^-16^**  0.39  **0.016**  0.37 | NA  - | |

*Supplementary table 4: Unadjusted and adjusted hazard ratios for risk to develop EDSS 8*

|  | Hazard ratio (95% CI) | p-value |
| --- | --- | --- |
| Genomic cluster  2  3 | 1.40 (0.99-1.98)  **1.45 (1.08-1.96)** | 0.059  **0.013** |
| Genomic cluster  2  3  Female sex | **1.45 (1.02-2.06)**  **1.42 (1.06-1.91)**  **0.67 (0.51-0.88)** | **0.037**  **0.020**  **0.0036** |
| Genomic cluster  2  3  Female sex  Age at onset | 1.41 (0.99-2.00)  **1.52 (1.12-2.04)**  **0.74 (0.56-0.97)**  **0.74 (0.56-0.97)** | 0.054  **0.0063**  **0.029**  **8*10^-11^** |
| Genomic cluster  2  3  Female sex  Age at onset  Disease modifying treatment  Only moderate efficacy  Only high efficacy  Switch moderate and high | **1.45 (1.02-2.06)**  **1.49 (1.11-2.01)**  **1.33 (0.58-0.99)**  **1.04 (1.03-1.06)**  **0.27 (0.13-0.55)**  0.90 (0.37-2.19)  0.63 (0.26-1.53) | **0.038**  **0.0087**  **0.041**  **1*10^-8^**  **0.00032**  0.81  0.31 |

*Supplementary table 5: Unadjusted and adjusted hazard ratios for risk to develop secondary progressive multiple sclerosis*

|  | Hazard ratio (95% CI) | p-value |
| --- | --- | --- |
| Genomic cluster  2  3 | 1.22 (0.98-1.51)  **1.25 (1.04-1.50)** | 0.076  **0.02** |
| Genomic cluster  2  3  Female sex | 1.24 (1.00-1.54)  **1.24 (1.025-1.49)**  **0.72 (0.60-0.85)** | 0.055  **0.026**  **0.00019** |
| Genomic cluster  2  3  Female sex  Age at onset | **1.27 (1.02-1.58)**  **1.31 (1.09-1.58)**  **0.75 (0.63-0.90)**  **1.04 (1.03-1.05)** | **0.033**  **0.0048**  **0.0016**  **3*10^-16^** |
| Genomic cluster  2  3  Female sex  Age at onset  Disease modifying treatment  Only moderate efficacy  Only high efficacy  Switch moderate and high | **1.27 (1.02-1.58)**  **1.30 (1.08-1.56)**  **0.75 (0.63-0.90)**  **1.04 (1.03-1.05)**  **0.80 (0.64-1.00)**  1.05 (0.69-1.60)  0.97 (0.68-1.38) | **0.03**  **0.006**  **0.0018**  **4*10^-16^**  **0.048**  0.82  0.84 |

*Supplementary Table 6: Results of multi-level modelling, estimates compared to genetic cluster 1*

| **Variable** | **Estimate (SE)** | **p-value** |
| --- | --- | --- |
| Genetic cluster 2 (onset) | 0.52 (0.43) | 0.23 |
| *Genetic cluster 2 (15 years)* | *0.38 (0.19)* | *0.049* |
| Genetic cluster 3 (onset) | -0.019 (0.36) | 0.96 |
| Genetic cluster 3 (15 years) | 0.18 (0.16) | 0.27 |

At disease onset, EDSS scores are similar between genomic cluster 1 versus 2 and 3 (resp. p=0.23 and 0.96). Patients in genomic cluster have a significantly higher increase in EDSS compared to cluster 1 (0.38 higher increase in EDSS/follow up year), whereas no difference was observed in cluster 3 versus cluster 1. All estimates are adjusted for use of DMT.

*Supplementary figure 1: Genomic clusters not associated with time to EDSS 4 in the Welsh cohort*

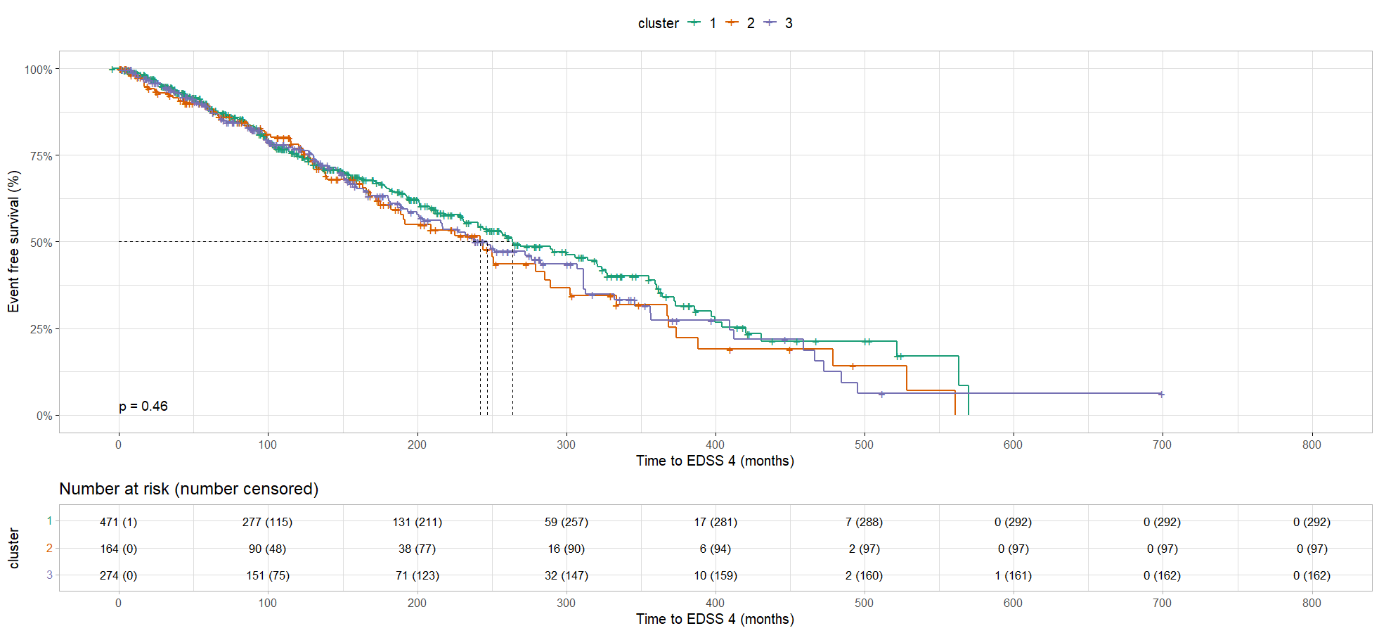

*Supplementary figure 2: Genetic clustering improves risk prediction of long-term disability compared to the number of relapses in the first 5 years after disease onset in Welsh patients*

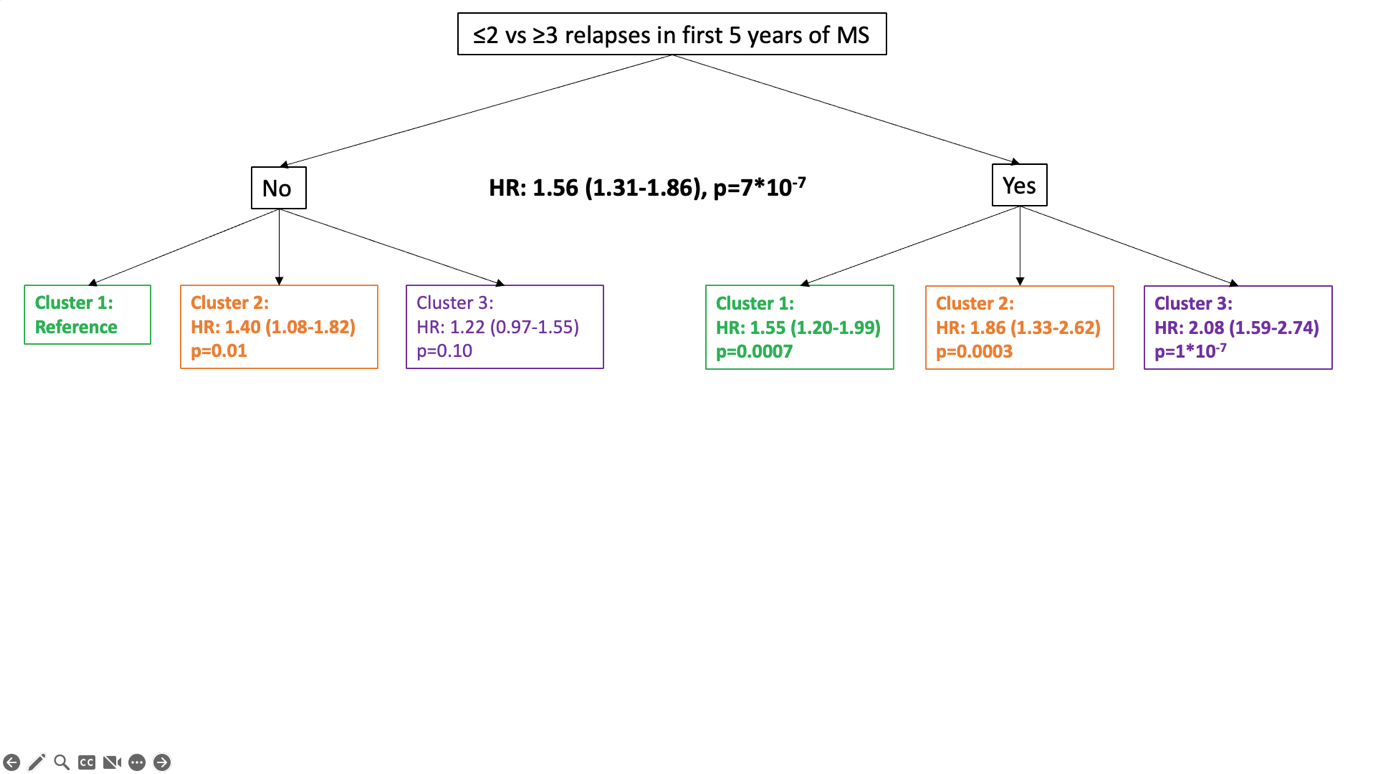
 No represents ≤2 relapses in first 5 years after disease onset

*Supplementary figure 3: MS DMT not affecting time to EDSS 4 in Welsh pwMS*

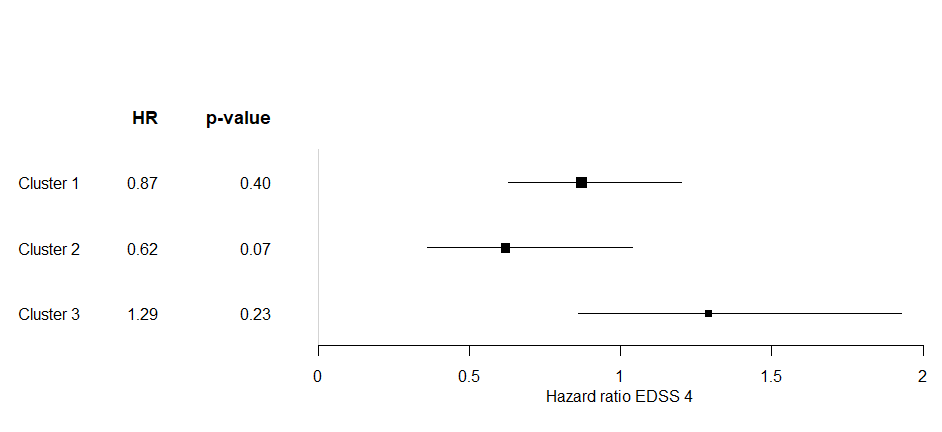
